# Family-related experiences, childhood trauma, and cognitive-emotional correlates across psychopathological domains in adolescent depression

**DOI:** 10.64898/2026.07.27.26358988

**Authors:** Ping Wang, Peijia Wang, Yaoyin Zhang, Xueyan Wang, Chao Li, Yulan Huang, Michael Maes

## Abstract

**Background:** Adolescent major depressive disorder (MDD) is heterogeneous, with diverse and frequently co-occurring psychopathological manifestations. Family-related experiences, childhood trauma, rumination, and psychological resilience are linked to adolescent mental health, but whether their associations are broadly shared or differ across clinical domains remains unclear. This study examined these associations within an integrative framework.

**Methods:** This cross-sectional study included 80 adolescents with MDD and 53 healthy controls. Family functioning, parenting, childhood trauma, rumination, psychological resilience, and four clinical domains comprising affective distress, suicidal ideation, the non-suicidal self-injury (NSSI) spectrum, and self-regulation difficulties were assessed using validated instruments. Partial least squares structural equation modeling examined multivariate associations. MDD-only and covariate-adjusted sensitivity analyses assessed robustness.

**Results:** The four clinical domains showed high reliability and generally acceptable discriminant validity. Family dysfunction was associated with maladaptive parenting and childhood trauma, and maladaptive parenting was associated with childhood trauma (β =.383–.600; all p <.001). Childhood trauma was associated with greater rumination (β =.625) and lower psychological resilience (β = −.405; both p <.001). Rumination and psychological resilience showed opposing associations with all four domains (rumination: β =.280–.559; resilience: β = −.399 to −.265; all p ≤.006). Family dysfunction, maladaptive parenting, and childhood trauma showed significant total indirect associations with all four domains (all p <.001). The model explained 53%– 85% of variance across the four domains. The MDD-only analysis largely preserved the overall structural pattern and showed broader direct associations of maladaptive parenting with the clinical domains. Covariate adjustment retained all principal associations except the association between psychological resilience and NSSI.

**Conclusions:** Psychopathology relevant to adolescent MDD was multidimensional, with related but distinguishable clinical domains sharing several psychosocial and cognitive-emotional correlates. The findings support examining domain-level psychopathology alongside diagnostic status and global depression severity.

**Plain Language Summary:** Adolescents with depression can show very different combinations of emotional distress, suicidal thoughts, self-injury, and problems with self-regulation. We asked whether several psychosocial and cognitive-emotional factors were linked to these different clinical problems in broadly similar ways, or whether their associations were more selective.

We studied 80 adolescents with major depressive disorder and 53 healthy adolescents. Family functioning, parenting, childhood trauma, rumination, and psychological resilience were examined together in relation to four areas of psychopathology: affective distress, suicidal ideation, non-suicidal self-injury, and self-regulation difficulties.

Rumination and lower psychological resilience were associated with greater difficulties across all four areas. Family dysfunction, maladaptive parenting, and childhood trauma were also related to each of the four areas through broader patterns of association. At the same time, the four clinical areas remained distinguishable rather than forming one undifferentiated severity score.

The same overall pattern was largely seen within adolescents with depression alone, although parenting showed broader direct associations in this group.

These findings suggest that adolescent depression may be better characterized by examining specific clinical domains alongside diagnosis and overall symptom severity.

## 1. Introduction

Depressive disorders are among the leading causes of disability worldwide and impose a substantial burden on individuals and society. ^1^ Adolescence is a particularly sensitive period marked by major biological, cognitive, emotional, and social changes, during which rates of depression rise markedly, especially among females. ^2–4^ Adolescent depression is associated with impaired academic functioning, family and peer relationships, and broader psychosocial development, as well as increased risks of recurrent depressive episodes, other psychiatric disorders, non-suicidal self-injury (NSSI), and suicidal behavior. ^5–6^ These consequences may extend beyond the acute phase and affect psychosocial functioning and mental health into adulthood. ^2,7–8^ A fuller characterization of depression during adolescence is therefore important for both research and clinical practice.

The clinical expression of depression during adolescence is highly variable. Adolescents may present with different combinations of affective symptoms, psychosomatic complaints, suicidal thoughts and behaviors, self-injury, and difficulties in self-regulation, ^2,9–10^ including among those who meet the same diagnostic criteria. Depressive symptoms also vary in severity, and the transition from relatively low symptom levels to clinically significant depression may not always form a clear categorical boundary. ^11^ Diagnostic status and overall depression severity therefore capture only part of the clinical variation relevant to adolescent mental health. Examining these manifestations as separate but related domains may provide a more informative account of clinical variation in adolescent depression.

Family experiences are widely recognized as an important developmental context in adolescent depression. ^12–13^ The family provides an enduring social environment in which emotional regulation, interpersonal expectations, and coping processes develop across childhood and adolescence. ^14^ Supportive family environments may facilitate adaptive coping and resilience, whereas family dysfunction may be associated with greater vulnerability to depression and related psychopathological problems. ^13,15^ Family dysfunction, including poor communication, emotional disengagement, chronic conflict, and ineffective problem-solving, has been associated with depressive symptoms and internalizing problems in adolescents. ^16–17^ Maladaptive parenting practices, such as rejection, psychological control, overprotection, and low emotional warmth, have likewise been associated with emotional and behavioral difficulties in youth. ^18–21^ Family dysfunction and maladaptive parenting thus represent related but distinct aspects of the family environment.

Exposure to childhood trauma is another important risk factor for mental health problems during childhood and adolescence and may have lasting consequences across the life course. ^22^ Adolescents with depression report higher levels of emotional abuse, emotional neglect, physical abuse, and other forms of childhood trauma than healthy peers. ^23–24^ Childhood trauma has been associated with a broad range of emotional and behavioral problems, including anxiety, substance use, disruptive behaviors, psychotic experiences, and suicidal thoughts and behaviors. ^25–28^ This diversity of outcomes raises a further question of how adolescents differ in their psychological responses to adverse experiences.

Rumination and psychological resilience are consistently associated with adolescent mental health and may help characterize individual differences in cognitive-emotional functioning. Rumination, characterized by repetitive and passive attention to negative emotions and their causes or consequences, has been associated with depressive symptoms, anxiety, suicidal ideation, and other forms of psychological distress. ^29–33^ Psychological resilience refers to the capacity to maintain or regain adaptive functioning in the face of stress or adversity and has been associated with more adaptive coping, lower emotional distress, and better psychological adjustment. ^31,34^ These constructs capture different aspects of cognitive-emotional functioning relevant to vulnerability and adaptation in adolescence. Rumination is generally associated with greater depressive vulnerability and more severe clinical manifestations, whereas resilience is associated with better psychological adjustment in the context of adversity. ^33–37^ Previous studies have established associations of family dysfunction, maladaptive parenting, childhood trauma, rumination, and psychological resilience with adolescent depression and related psychopathology. A further question is whether these factors show broadly shared patterns of association across the heterogeneous clinical manifestations of adolescent depression, or whether some associations vary across different aspects of the clinical presentation. Examining them within the same framework allows shared associations across clinical manifestations to be distinguished from more selective patterns while accounting for relationships among the factors themselves.

The present study therefore examined family dysfunction, maladaptive parenting, childhood trauma, rumination, and psychological resilience together within an integrative model. Clinical variation was represented by four prespecified domains encompassing affective distress, suicidal ideation, the NSSI spectrum, and self-regulation difficulties, allowing the same set of factors to be examined across different manifestations of psychopathology. We first examined the model across adolescents with MDD and healthy controls to characterize associations across a broad range of psychopathological variation. We then examined the model within adolescents with MDD to assess how these associations were expressed across clinical variation within the disorder itself.

## 2. Methods

### 2.1 Participants and Study Design

The Ethics Committee of the University of Electronic Science and Technology of China approved the study protocol (Approval No. 31682). For participants under 18 years, written informed consent was obtained from a parent or legal guardian, alongside written informed assent from the adolescents. Participants aged 18 years provided independent written informed consent.

This cross-sectional study enrolled adolescents with MDD from the inpatient unit of the Sichuan Provincial Center for Mental Health between December 2024 and January 2026. Diagnoses were established by experienced psychiatrists using DSM-5 criteria. Eligible patients met the following inclusion criteria: (1) age 12–18 years; (2) right-handedness; and (3) a 24-item Hamilton Depression Rating Scale (HAMD-24) score >17 at admission. Exclusion criteria included: (1) neurological disorders, significant head trauma, or severe medical conditions (e.g., diabetes mellitus or active infectious diseases); (2) comorbid psychiatric disorders, including bipolar disorder, schizophrenia spectrum disorders, autism spectrum disorder, intellectual disability, conduct disorder, obsessive-compulsive disorder, or other pervasive developmental disorders; (3) history of psychoactive substance abuse, except nicotine dependence.

Healthy controls, matched on age and sex frequency distributions, were recruited from the community and via social media. Controls were excluded if they had any current or lifetime DSM-5 psychiatric disorder, neurological condition, or severe medical illness. A total of 148 adolescents were assessed for eligibility. After exclusion of individuals due to ineligibility, refusal, or incomplete data, 133 participants were included in the final analysis. Participant flow is shown in Figure 1, and baseline characteristics are presented in Table 1.

**Figure 1:**
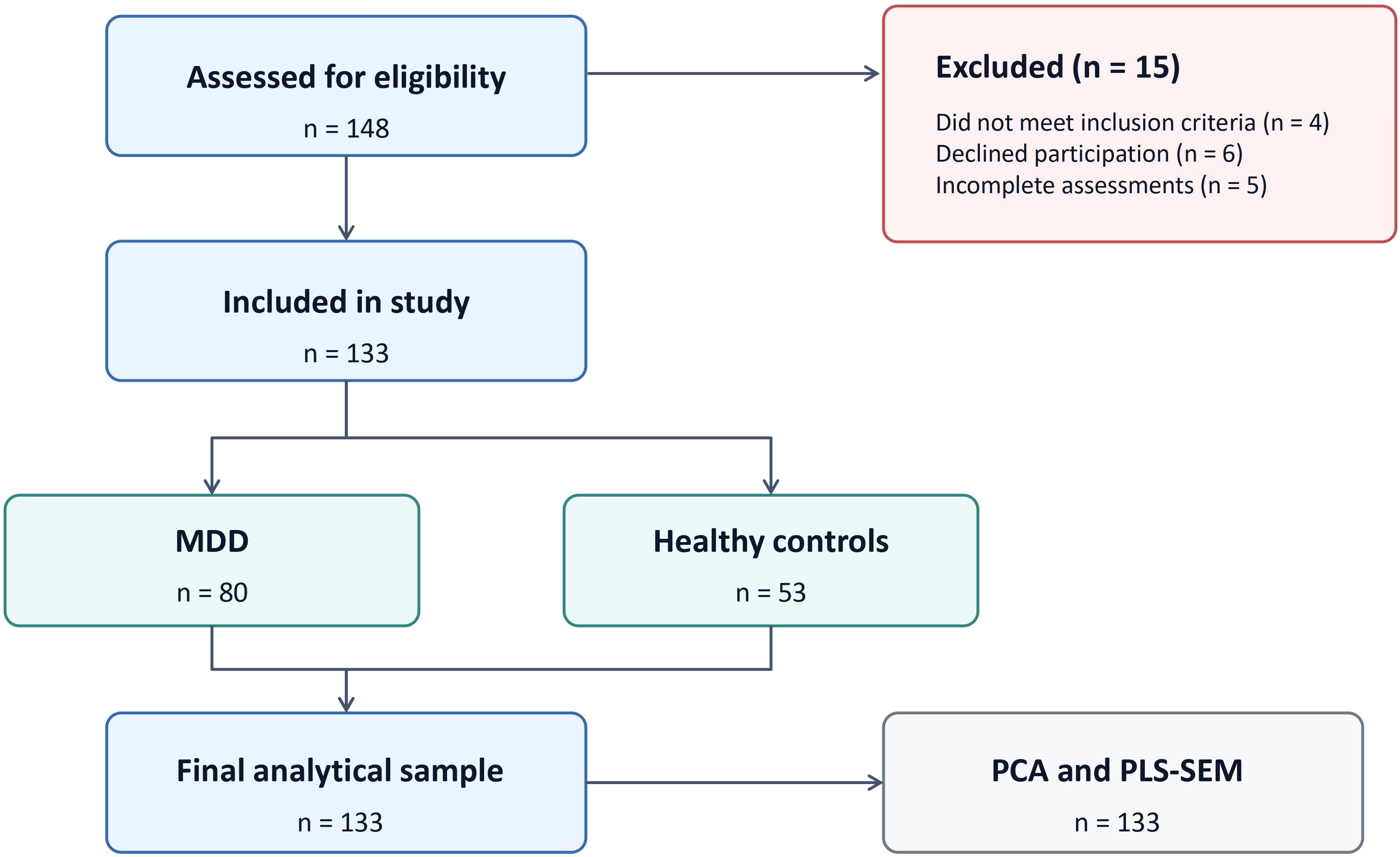
Participant recruitment and study flow. Of 148 adolescents assessed for eligibility, 133 met study criteria and were included in the final analyses, comprising 80 adolescents with major depressive disorder (MDD) and 53 healthy controls (HC).

**Table 1:** Comparison of demographic, clinical, psychosocial, and family characteristics between adolescents with major depressive disorder and healthy controls.

| Characteristic | HC (n=53) | MDD (n=80) | p-value |
| --- | --- | --- | --- |
| **Sociodemographic characteristics** |  |  |  |
| Age, y | 16 [13.5–17] | 15 [13.25–17] | 0.885 |
| BMI, kg/m<sup>2</sup> | 20.0 [17.3–21.4] | 19.5 [17.7–22.3] | 0.834 |
| Female sex, n (%) | 29 (54.7) | 52 (65.0) | 0.234 |
| Current school grade | 10th [7th–10th] | 10th [8th–11th] | 0.474 |
| **Family and academic environment** |  |  |  |
| Academic performance, n (%) |  |  | 0.893 |
| High grades | 14 (26.4) | 22 (27.5) |  |
| Middle grades | 33 (62.3) | 47 (58.8) |  |
| Lower grades | 6 (11.3) | 11 (13.8) |  |
| Academic pressure, n (%) |  |  | <0.001 |
| Low | 24 (45.3) | 7 (8.8) |  |
| Middle | 26 (49.1) | 31 (38.8) |  |
| High | 3 (5.7) | 42 (52.5) |  |
| Paternal education level, n (%) |  |  | <0.001 |
| Primary school or below | 5 (9.4) | 13 (16.3) |  |
| Middle school | 7 (13.2) | 29 (36.3) |  |
| High school | 6 (11.3) | 25 (31.3) |  |
| College | 35 (66.0) | 13 (16.3) |  |
| Maternal education level, n (%) |  |  | <0.001 |
| Primary school or below | 4 (7.5) | 15 (18.8) |  |
| Middle school | 12 (22.6) | 39 (48.8) |  |
| High school | 8 (15.1) | 16 (20.0) |  |
| College | 29 (54.7) | 10 (12.5) |  |
| Parental marital status, n (%) |  |  | 0.003 |
| Married/living together | 50 (94.3) | 59 (73.8) |  |
| Separated family | 3 (5.7) | 21 (26.3) |  |
| Primary caregiver, n (%) |  |  | <0.001 |
| Both parents | 47 (88.7) | 36 (45.0) |  |
| Single parent | 6 (11.3) | 35 (43.8) |  |
| Grandparents/others | 0 (0.0) | 9 (11.3) |  |
| Siblings, n (%) |  |  | 0.107 |
| No | 14 (26.4) | 32 (40.0) |  |
| Yes | 39 (73.6) | 48 (60.0) |  |
| **Psychopathology** |  |  |  |
| Depression (CDI) | 11 [6–16] | 34 [27.25–40.75] | <0.001 |
| Anxiety (SCARED) | 16 [10.5–22.5] | 55.5 [46–65] | <0.001 |
| Suicidal ideation (Beck) | 2 [1–8] | 26 [21–30] | <0.001 |
| Self-Regulation Difficulties (SNAP) | 12 [4–22.5] | 33 [21.3–46.5] | <0.001 |
| Resilience (CD-RISC) | 61 [49.5–71] | 32.5 [21–40] | <0.001 |
| Rumination (RRS) | 19 [11–28] | 46 [35.25–55] | <0.001 |
| **Family-related experiences and childhood trauma** |  |  |  |
| Childhood trauma (CTQ) | 11 [7.25–17] | 28 [19.25–37.5] | <0.001 |
| Maladaptive parenting (PCRT) | 67 [58–85.5] | 87 [71–106] | 0.001 |
| Family dysfunction (FAD) | 128 [111–144.5] | 152 [137.75–161] | <0.001 |

Notes. Values are median [IQR] unless otherwise indicated. Group comparisons used Mann–Whitney U or chi-square tests as appropriate. Missing data were handled using pairwise deletion. Higher scores indicate greater severity for CTQ, PCRT, and FAD.

### 2.2 Clinical and Psychosocial Assessments

Demographic and clinical data were obtained from medical records and verified through semi-structured clinical interviews. Clinician-rated depressive and anxiety symptom severities were evaluated using the validated Chinese versions of the HAMD-24 ^38^ and the Hamilton Anxiety Rating Scale (HAMA-14). ^39^

Upon enrollment, participants completed a standardized assessment battery using Chinese versions of established clinician-administered and self-report instruments. Depressive and anxiety symptoms were assessed using the Children’s Depression Inventory (CDI) ^40^ and the Screen for Child Anxiety Related Emotional Disorders (SCARED), ^41^ respectively. Attentional and behavioral difficulties were measured using the Swanson, Nolan, and Pelham Rating Scale (SNAP-IV). ^42^ Ruminative response style and psychological resilience were assessed using the Ruminative Responses Scale (RRS) ^43^ and the Connor–Davidson Resilience Scale (CD-RISC), ^44^ respectively. Suicidal ideation and self-injurious behaviors were assessed using the Columbia– Suicide Severity Rating Scale (C-SSRS), ^45^ the Beck Scale for Suicide Ideation (BSSI), ^46^ and the Ottawa Self-Injury Inventory (OSI). ^47^ Family-related experiences were assessed using the Family Assessment Device (FAD), ^48^ the Parent–Child Relationship Diagnostic Test (PCRT), ^49^ while childhood trauma was assessed using the Childhood Trauma Questionnaire–Short Form (CTQ-SF). ^50^

### 2.3 Theoretical Domain Specification and Preliminary Dimensionality Assessment

The psychosocial and clinical domains were specified a priori based on the theoretical framework, the constructs assessed by the original instruments, and their established scale or subscale structures. Principal component analysis (PCA) was conducted separately within each prespecified domain to evaluate whether the theoretically defined indicator sets exhibited a dominant underlying dimension consistent with their conceptual structure. PCA was used only for preliminary dimensionality assessment and for deriving summary scores for descriptive and bivariate analyses; the PLS-SEM model used the prespecified observed scale or subscale indicators.

Within each domain, dimensionality was assessed using eigenvalues (>1.0), scree plots, variance explained by the first component, indicator loadings, and conceptual interpretability. A one-component solution was retained when these criteria consistently supported a dominant dimension.

For the childhood trauma domain, the sexual abuse subscale was excluded because it showed a low loading in the preliminary dimensionality assessment.

For descriptive and bivariate analyses, first-component scores were used as summary measures of each domain. Nine domains were evaluated: family dysfunction (FAD subscales), maladaptive parenting (PCRT subscales), childhood trauma (CTQ subscales), rumination (RRS dimensions), psychological resilience (CD-RISC dimensions), affective distress (CDI and SCARED scores), suicidal ideation (BSSI and C-SSRS indicators), NSSI spectrum (OSI behavioral indicators), and self-regulation difficulties (SNAP-IV dimensions). The component matrices, variance explained, and indicator loadings for the prespecified domains are presented in Table 2.

**Table 2:** Principal component analysis of the prespecified psychosocial and psychopathological domains.

| Construct | Indicators | KMO | Bartlett's p | Variance explained (%) | PCA loadings |
| --- | --- | --- | --- | --- | --- |
| Family dysfunction | PS, CM, RL, AR, AI, BC, GF | 0.883 | <0.001 | 67.67 | 0.735–0.915 |
| Maladaptive parenting | Rejection, Control, Protection, Inconsistency | 0.740 | <0.001 | 65.57 | 0.708–0.917 |
| Childhood trauma | PN, EN, EA, PA | 0.683 | <0.001 | 59.84 | 0.647 – 0.906 |
| Rumination | SR, B, RP | 0.767 | <0.001 | 88.92 | 0.934–0.948 |
| Psychological resilience | Tenacity, Strength, Optimism | 0.767 | <0.001 | 88.64 | 0.936–0.948 |
| Affective distress | CDI, SCARED | 0.500 | <0.001 | 95.47 | 0.977 |
| Suicidal ideation | Beck-LW, Beck-AL, SI severity | 0.670 | <0.001 | 85.14 | 0.896–0.964 |
| Self-regulation difficulties | SNAP-AD, SNAP-HI, SNAP-ODD | 0.738 | <0.001 | 77.86 | 0.883–0.898 |
| NSSI spectrum | Frequency, methods, locations | 0.768 | <0.001 | 76.82 | 0.859–0.889 |

*Notes. PCA component scores were used for descriptive and bivariate analyses, whereas the PLS-SEM used the prespecified scale or subscale indicators. PS = Problem Solving; CM = Communication; RL = Roles; AR = Affective Responsiveness; AI = Affective Involvement; BC = Behavior Control; GF = General Functioning. PN = Physical Neglect; EN = Emotional Neglect; EA = Emotional Abuse; PA = Physical Abuse. SR = Symptom Rumination; B = Brooding; RP = Reflective Pondering. Beck-LW = Beck Scale for Suicidal Ideation (last week); Beck-AL = Beck Scale for Suicidal Ideation (lifetime). SNAP = Swanson, Nolan, and Pelham Questionnaire; NSSI = non-suicidal self-injury.*

### 2.4 Statistical Analyses

Demographic, clinical, and psychosocial characteristics were compared between the MDD and healthy control groups using univariable analyses. The distributions of continuous variables were assessed using the Shapiro-Wilk test. Mann-Whitney U tests were used for non-normally distributed continuous variables, whereas χ² tests or Fisher’s exact tests were used for categorical variables, as appropriate. The proportion of missing data was low (<5% for all variables). Pairwise available-case analysis was used for descriptive and bivariate analyses. Detailed clinical characteristics specific to the MDD group are reported in Supplementary Table S1.

Bivariate associations among domain summary scores were examined using Spearman rank-order correlations.

Structural associations were analyzed using PLS-SEM. The model specified relationships among theoretically prespecified constructs, with observed scale or subscale scores serving as indicators. Statistical inference was based on bootstrapping with 5,000 resamples using two-tailed tests, with statistical significance set at p <.05. Model evaluation included indicator loadings, internal consistency reliability, convergent validity assessed using average variance extracted (AVE), discriminant validity assessed using the heterotrait-monotrait ratio (HTMT), standardized root mean square residual (SRMR), coefficients of determination (R²), inner model collinearity diagnostics, PLSpredict-derived Q²_predict_ values, and out-of-sample predictive performance evaluated using the cross-validated predictive ability test (CVPAT). HTMT point estimates below.90 were treated as the primary heuristic for discriminant validity, with bootstrap confidence intervals also inspected.

A staged analytical approach was adopted. The primary analysis was conducted in the combined HC+MDD sample to capture associations across a broad spectrum of psychosocial and clinical variation. Sensitivity analyses were subsequently performed in the MDD-only subsample (n=80) to assess whether the main structural pattern was retained within the clinical population.

To further evaluate the robustness of the observed structural associations, an auxiliary covariate-adjusted analysis was conducted using individual-level latent variable scores extracted from the final PLS-SEM model. Each structural equation was re-estimated using standardized ordinary least squares regression, adjusting for paternal education, maternal education, primary caregiver structure, parental marital status, and academic pressure. These covariates were selected because they differed between groups and may be associated with family context and clinical domains. Categorical variables were dummy-coded. Statistical uncertainty was assessed using 5,000 bootstrap resamples. This analysis was intended as a supplementary robustness check rather than a replacement for the primary PLS-SEM model.

Primary analyses were conducted using IBM SPSS Statistics (version 31; IBM Corp.) and SmartPLS 4 (SmartPLS GmbH). The regression-based covariate-adjusted sensitivity analysis was conducted using Python with NumPy and pandas.

## 3. Results

### 3.1 Demographic and Clinical Characteristics

The final analyzed cohort comprised 133 adolescents (80 with MDD and 53 healthy controls). The groups did not differ in age, sex, BMI, school grade, academic performance, or sibling status (all p >.05). In contrast, group differences were observed in academic pressure, parental education, caregiver structure, and parental marital status (all p <.01).

Compared with healthy controls, adolescents with MDD showed higher scores across clinical and psychosocial measures, including depressive and anxiety symptoms, suicidal ideation, self-regulation difficulties, rumination, childhood trauma, maladaptive parenting, and family dysfunction (all p ≤.001). Resilience scores were lower in the MDD group (p <.001; Table 1).

### 3.2 Composite Domain Construction

Within each prespecified domain, PCA supported a single-component solution (Table 2). KMO values ranged from 0.500 to 0.883, and Bartlett tests were significant across all domains (p <.001). The first principal components explained 59.84%–95.47% of the variance, with item loadings ranging from 0.647 to 0.977. Component scores were retained for descriptive and bivariate analyses.

### 3.3 Bivariate Correlations Among Composite Domains

Spearman correlations in the combined sample (N = 133) showed significant associations across the composite domains (all p <.001; Table S2 and Figure 2). Family dysfunction and maladaptive parenting were strongly correlated with childhood trauma. Childhood trauma was correlated with rumination, psychological resilience, and all clinical domains. The strongest positive correlation was observed between rumination and affective distress (ρ = 0.851), whereas the strongest negative correlation was observed between psychological resilience and affective distress (ρ = −0.821).

**Figure 2:**
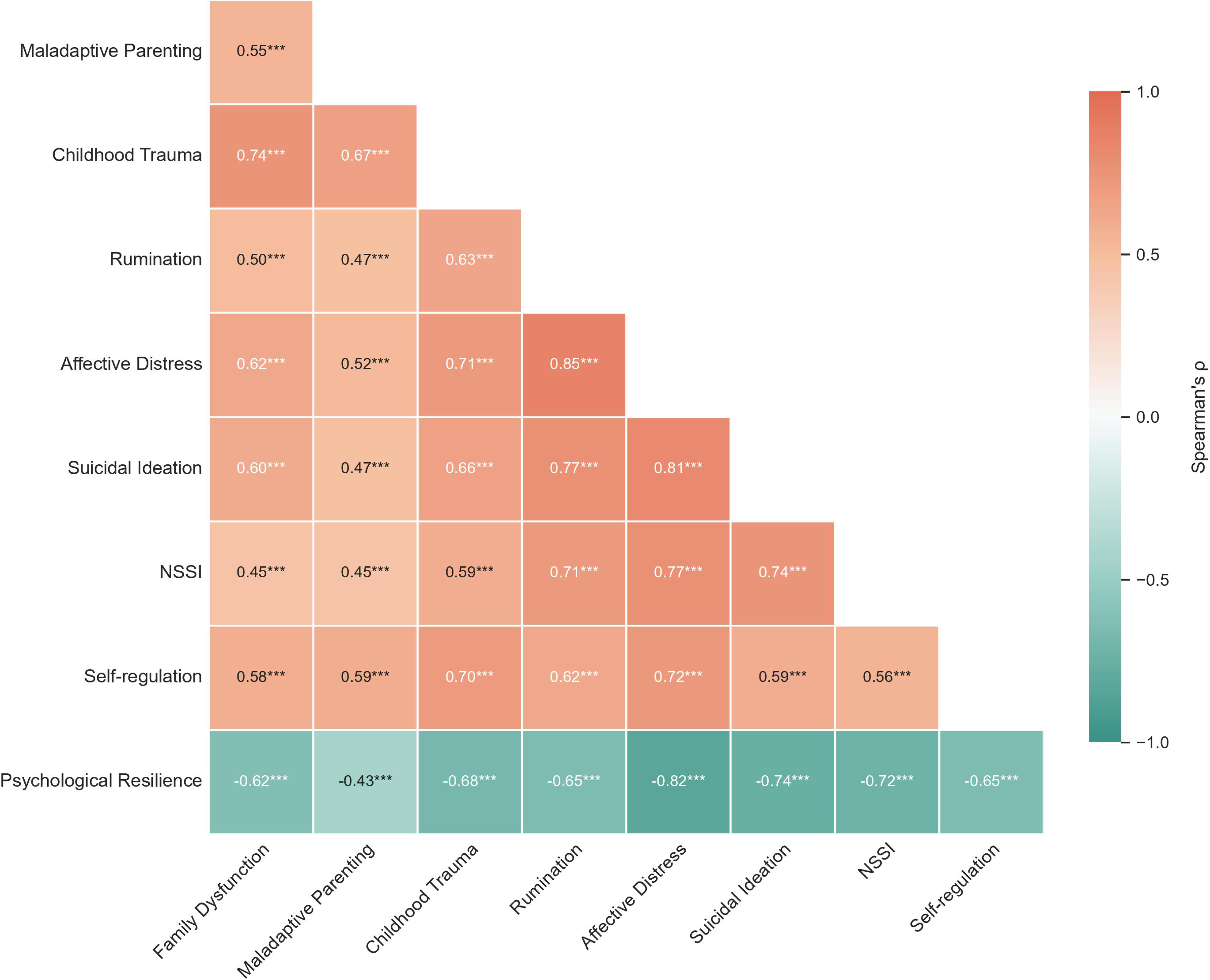
Spearman correlations among psychosocial, cognitive-emotional, and psychopathological domain scores in the combined sample. Cells display Spearman’s rank correlation coefficients (ρ). Warmer colors indicate positive correlations, whereas cooler colors indicate negative correlations. ***p <.001, two-tailed. NSSI, non-suicidal self-injury.

### 3.4 PLS-SEM Model Evaluation and Structural Associations

The measurement model demonstrated satisfactory internal consistency and convergent validity across all multi-indicator constructs, with Cronbach’s α ranging from 0.768 to 0.953, composite reliability from 0.852 to 0.977, and average variance extracted (AVE) from 0.594 to 0.955. HTMT point estimates were below.90 for all but two construct pairs: rumination–affective distress (.909) and suicidal ideation–affective distress (.901). All bootstrap 95% confidence intervals had upper bounds below 1.00 (Table S3). Among the four clinical domains, HTMT values ranged from.689 to.901, indicating generally acceptable but not complete separation. Explained variance (R²) ranged from 0.360 for maladaptive parenting to 0.854 for affective distress. Q²_predict_ values were positive for all endogenous variables. Inner model collinearity diagnostics indicated no problematic multicollinearity (maximum VIF = 2.06). The saturated-model and estimated-model SRMR values were.082 and.099, respectively. For affective distress, the reported CVPAT comparison favored the PLS-SEM predictions over the linear benchmark (p <.001; Table S4).

The final structural model is presented in Figure 3, and standardized direct associations are summarized in Table 3. Family dysfunction was positively associated with maladaptive parenting (β = 0.600, p <.001) and childhood trauma (β = 0.478, p <.001), whereas maladaptive parenting was additionally associated with childhood trauma (β = 0.383, p <.001).

**Figure 3:**
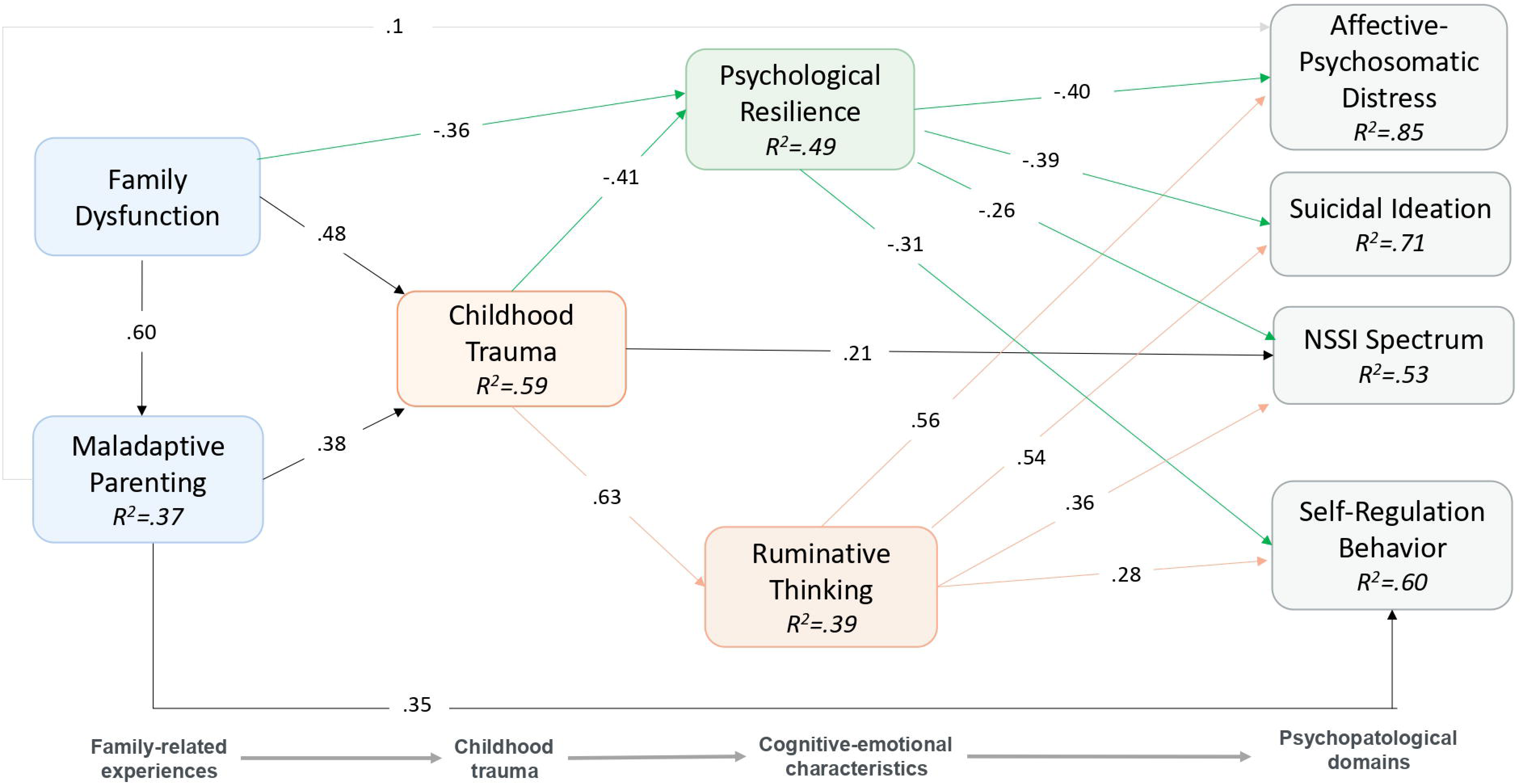
Final PLS-SEM model in the combined sample. The model depicts associations among family dysfunction, maladaptive parenting, childhood trauma, rumination, psychological resilience, and four psychopathological domains. Values represent standardized path coefficients (β). Only statistically significant structural paths (p <.05) are displayed. Numbers within endogenous constructs indicate explained variance (R²).

**Table 3:** Standardized direct associations in the PLS-SEM model for the combined sample.

| Path | $\beta$ | SE | t | p |
| --- | --- | --- | --- | --- |
| ***Family-related structural associations*** |  |  |  |  |
| Family Dysfunction → Maladaptive Parenting | .600 | .049 | 12.303 | <.001 |
| Family Dysfunction → Childhood Trauma | .478 | .058 | 8.298 | <.001 |
| Maladaptive Parenting → Childhood Trauma | .383 | .059 | 6.551 | <.001 |
| ***Associations with rumination and resilience*** |  |  |  |  |
| Childhood Trauma → Rumination | .625 | .051 | 12.277 | <.001 |
| Childhood Trauma → Psychological Resilience | -.405 | .100 | 4.051 | <.001 |
| Family Dysfunction → Psychological Resilience | -.355 | .104 | 3.398 | .001 |
| ***Associations of rumination and resilience with psychopathological domains*** |  |  |  |  |
| Rumination → Affective distress | .559 | .048 | 11.613 | <.001 |
| Rumination → Suicidal Ideation | .535 | .060 | 8.933 | <.001 |
| Rumination → NSSI Spectrum | .356 | .087 | 4.098 | <.001 |
| Rumination → Self-regulation difficulties | .280 | .085 | 3.297 | .001 |
| Psychological Resilience → Affective distress | -.399 | .046 | 8.708 | <.001 |
| Psychological Resilience → Suicidal Ideation | -.394 | .055 | 7.137 | <.001 |
| Psychological Resilience → Self-regulation difficulties | -.308 | .074 | 4.172 | <.001 |
| Psychological Resilience → NSSI Spectrum | -.265 | .096 | 2.759 | .006 |
| ***Additional direct associations with psychopathological domains*** |  |  |  |  |
| Childhood Trauma → NSSI Spectrum | .214 | .094 | 2.273 | .023 |
| Maladaptive Parenting → Self-regulation difficulties | .352 | .068 | 5.193 | <.001 |
| Maladaptive Parenting → Affective distress | .098 | .040 | 2.451 | .014 |

Childhood trauma was positively associated with rumination (β = 0.625, p <.001) and negatively associated with psychological resilience (β = –0.405, p <.001). Family dysfunction also showed an additional direct association with lower psychological resilience (β = –0.355, p =.001).

Rumination was positively associated with all four clinical domains, including affective distress, suicidal ideation, the NSSI spectrum, and self-regulation difficulties (all p ≤.001). Psychological resilience showed inverse associations with all four clinical domains (all p ≤.01). Additional direct associations were observed from childhood trauma to the NSSI spectrum and from maladaptive parenting to affective distress and self-regulation difficulties.

The final model explained 36.0% of the variance in maladaptive parenting, 59.6% in childhood trauma, 39.1% in rumination, 49.4% in psychological resilience, 85.4% in affective distress, 70.9% in suicidal ideation, 53.4% in the NSSI spectrum, and 60.1% in self-regulation difficulties (Figure 3).

Significant total indirect associations are summarized in Table 4. Family dysfunction and maladaptive parenting showed significant indirect associations with all four psychopathological domains. Childhood trauma also showed significant indirect associations with affective distress, suicidal ideation, self-regulation difficulties, and the NSSI spectrum.

**Table 4:** Standardized total indirect associations in the PLS-SEM model for the combined.

| Indirect association | $\beta$ | SE | t | p |
| --- | --- | --- | --- | --- |
| Indirect associations of Family Dysfunction |  |  |  |  |
| Family Dysfunction → Affective distress | .563 | .045 | 12.584 | <.001 |
| Family Dysfunction → Suicidal Ideation | .490 | .043 | 11.476 | <.001 |
| Family Dysfunction → NSSI Spectrum | .480 | .048 | 10.098 | <.001 |
| Family Dysfunction → Self-regulation difficulties | .533 | .045 | 11.903 | <.001 |
| Indirect associations of Maladaptive Parenting |  |  |  |  |
| Maladaptive Parenting → Affective distress | .196 | .037 | 5.234 | <.001 |
| Maladaptive Parenting → Suicidal Ideation | .189 | .038 | 5.020 | <.001 |
| Maladaptive Parenting → NSSI Spectrum | .209 | .044 | 4.787 | <.001 |
| Maladaptive Parenting → Self-regulation difficulties | .115 | .029 | 3.932 | <.001 |
| Indirect associations of Childhood Trauma |  |  |  |  |
| Childhood Trauma → Affective distress | .511 | .053 | 9.666 | <.001 |
| Childhood Trauma → Suicidal Ideation | .494 | .055 | 9.040 | <.001 |
| Childhood Trauma → NSSI Spectrum | .330 | .058 | 5.694 | <.001 |
| Childhood Trauma → Self-regulation difficulties | .300 | .058 | 5.143 | <.001 |

### 3.5 Sensitivity Analysis in the MDD-Only Sample

The overall structural pattern was largely preserved in the MDD-only sample (Figure 4). Many of the principal structural associations among family dysfunction, maladaptive parenting, childhood trauma, rumination, psychological resilience, and the four clinical domains remained statistically significant (Table S6).

**Figure 4:**
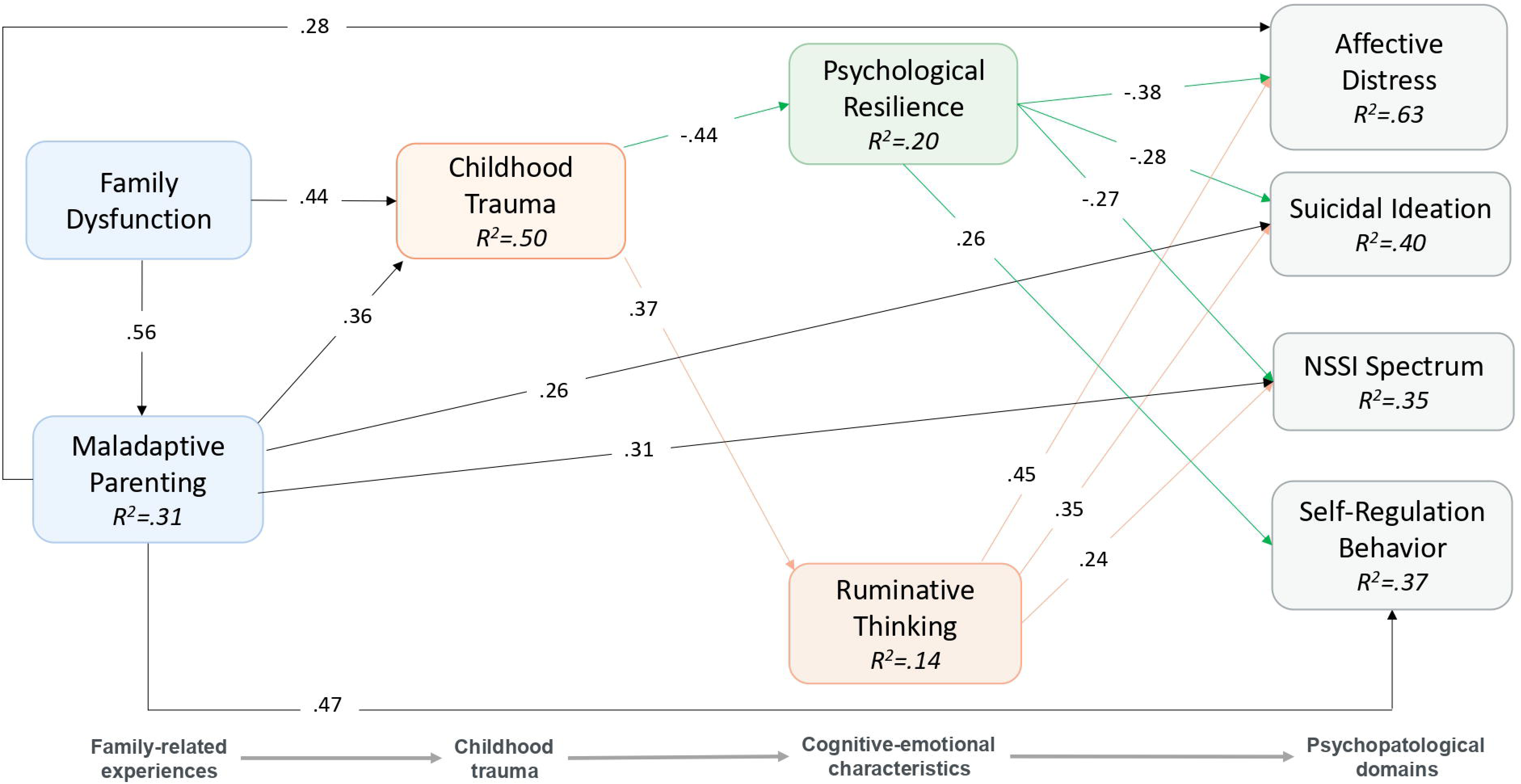
PLS-SEM model in the MDD-only sample. Sensitivity analysis restricted to adolescents with MDD. Values represent standardized path coefficients (β). Only statistically significant structural paths (p <.05) are displayed. Numbers within endogenous constructs indicate explained variance (R²).

Several coefficients were smaller in the MDD-only model (Table S6), including the associations from childhood trauma to rumination and from rumination and psychological resilience to suicidal ideation. Maladaptive parenting remained directly associated with affective distress and self-regulation difficulties, and additional direct associations with suicidal ideation and the NSSI spectrum were observed. Overall, the main pattern of structural associations remained similar.

### 3.6 Covariate-Adjusted Sensitivity Analysis

Adjustment for paternal education, maternal education, primary caregiver structure, parental marital status, and academic pressure did not materially alter the overall structural pattern identified in the primary PLS-SEM model (Table S7). The direction and statistical significance of nearly all substantive structural associations were preserved after adjustment.

Several path coefficients were modestly attenuated following covariate adjustment, most notably the associations from childhood trauma to rumination and psychological resilience, and from rumination and psychological resilience to suicidal ideation. The direct associations of maladaptive parenting with affective distress and self-regulation difficulties remained statistically significant after adjustment. The association between psychological resilience and the NSSI spectrum was no longer statistically significant after covariate adjustment (p =.067).

## 4. Discussion

The present study examined whether family-related experiences, childhood trauma, rumination, and psychological resilience showed broadly shared or differing associations across multiple clinical domains relevant to adolescent depression. Three findings are particularly informative. Greater rumination was associated with higher levels of affective distress, suicidal ideation, the NSSI spectrum, and self-regulation difficulties. Lower psychological resilience showed a similarly broad pattern, with inverse associations across all four domains. Family dysfunction, maladaptive parenting, and childhood trauma also each showed significant total indirect associations with every clinical domain. Despite these shared associations, the measurement model provided overall support for separation of the four domains, indicating that their common psychosocial and cognitive-emotional correlates did not simply reflect a single undifferentiated clinical dimension. Finally, the overall structural pattern was largely preserved within adolescents with MDD, while maladaptive parenting showed broader direct associations with the clinical domains in this restricted sample. Together, these findings suggest substantial commonality across the clinical manifestations of adolescent depression, alongside some differences in how individual psychosocial factors relate to particular domains.

Family dysfunction, maladaptive parenting, and childhood trauma showed close associations within the structural model, consistent with previous evidence linking family functioning, parenting experiences, and childhood maltreatment in adolescent mental health. ^13,31^ At the same time, each construct represented a different aspect of adolescents’ psychosocial experience. Family dysfunction reflects broader patterns of communication, problem-solving, emotional involvement, and family organization. Maladaptive parenting reflects more specific patterns of parent–child interaction, while childhood trauma captures experiences of abuse and neglect. Keeping these constructs separate was therefore important for examining whether related psychosocial experiences showed similar or different patterns of association across the four clinical domains.

Rumination and psychological resilience showed broad associations across the four clinical domains. Greater rumination was associated with higher affective distress, suicidal ideation, the NSSI spectrum, and self-regulation difficulties, while higher psychological resilience was associated with lower levels of all four domains. Previous studies have linked rumination and resilience to depression, suicidality, and self-injury in adolescents. ^29,31,34–37^ Examining the two characteristics within the same model showed that both were relevant across several clinical manifestations and were not confined to a single domain. Their associations with family-related experiences and childhood trauma were not identical. Childhood trauma was associated with both greater rumination and lower resilience, while family dysfunction showed an additional association with lower resilience. Taken together, these findings suggest that rumination and resilience contribute to a shared cognitive-emotional pattern across the clinical domains, while retaining different associations with the psychosocial factors examined in the model.

The four clinical domains showed both commonality and differentiation. Affective distress, suicidal ideation, the NSSI spectrum, and self-regulation difficulties were substantially correlated, and the measurement model generally supported their discriminant validity. Their broad associations with rumination and psychological resilience, together with the indirect associations of family dysfunction, maladaptive parenting, and childhood trauma across all four domains, indicate that these manifestations share several psychosocial and cognitive-emotional correlates. Additional direct associations were observed for particular domains, indicating that the four domains were not interchangeable expressions of a single clinical severity dimension. These findings should not be interpreted as evidence for discrete subtypes of adolescent depression. They instead support examining these manifestations as related but distinguishable domains alongside diagnostic status and global depression severity. In clinical practice, assessing suicidal ideation, NSSI, and self-regulation difficulties separately from overall affective symptom burden may help identify which problems require closer attention in an individual adolescent and support more differentiated clinical monitoring and management.

The MDD-only analysis largely preserved the overall structural pattern observed in the combined sample. Similar associations were evident among adolescents who already met diagnostic criteria for MDD, where the analysis focused on variation in clinical expression within the disorder. Several coefficients were smaller in the MDD-only model, while maladaptive parenting showed direct associations with all four clinical domains. These differences should be interpreted cautiously because restricting the analysis to the MDD group reduced both the sample size and the range of psychopathological variation. Even so, the broader associations of maladaptive parenting suggest that variation in parent–child relationships may remain clinically relevant to differences in presentation among adolescents with MDD. The covariate-adjusted analyses also retained nearly all principal associations, although the association between psychological resilience and the NSSI spectrum was no longer statistically significant. Overall, these sensitivity analyses support the stability of the main pattern while indicating that individual associations were not equally robust across analytical conditions.

### 4.1 Limitations

Several limitations should be considered when interpreting these findings. First, the cross-sectional design does not establish temporal ordering or causality. The directional paths and indirect associations in the PLS-SEM model should therefore be interpreted as theory-informed statistical associations and not as evidence of temporal mediation or developmental mechanisms. Longitudinal studies are needed to determine the temporal ordering and developmental stability of these relationships.

Second, the primary model combined healthy controls and adolescents with MDD to capture a broad range of psychopathological variation. This design increases variation in the measured constructs, but some associations may partly reflect systematic differences between the two groups. The largely preserved structural pattern in the MDD-only analysis reduces this concern, although it does not eliminate it.

Third, the four clinical domains were operationalized using the measures available in the present cohort. Although the domains showed high reliability and generally acceptable discriminant validity, they should not be regarded as an exhaustive or definitive taxonomy of adolescent psychopathology. Replication using independent samples and alternative measures will be important for determining the stability and generalizability of this multidomain structure. The present model was also restricted to psychosocial and cognitive-emotional factors and did not encompass biological, treatment-related, or other contextual influences.

Fourth, family-related experiences, childhood trauma, rumination, and psychological resilience were assessed primarily through adolescent self-report. These measures may be influenced by recall bias, current emotional state, and shared method variance. Future studies incorporating parent or clinician reports, behavioral measures, and prospective assessments may provide stronger multimethod evaluation of these associations.

Finally, participants were recruited from a single inpatient center in China, and the sample size was modest, particularly for the MDD-only analysis. The differences observed between the combined and MDD-only models should therefore be interpreted cautiously, particularly where they are based on changes in the magnitude or statistical significance of individual paths rather than formal between-model comparisons. Independent replication in larger clinical and community samples and across different cultural settings will be needed to evaluate the generalizability of the findings.

## 5. Conclusion

The present study provides an integrated account of how family-related experiences, childhood trauma, rumination, and psychological resilience are associated with the multidimensional clinical expression of adolescent depression. Family dysfunction, maladaptive parenting, and childhood trauma were interrelated, while rumination and psychological resilience showed broad and opposing associations with all four clinical domains. Family dysfunction, maladaptive parenting, and childhood trauma also showed indirect associations across the four domains. The domains were related and showed generally acceptable discriminant validity, supporting their consideration as distinguishable domains relevant to the clinical expression of adolescent MDD. Future research may benefit from incorporating quantitative measures of affective distress, suicidal ideation, the NSSI spectrum, and self-regulation difficulties alongside diagnostic status and global depression severity. Longitudinal and multimodal studies are needed to determine whether these domains differ in their developmental trajectories, biological correlates, and responses to preventive or therapeutic interventions.

## 6. Declarations

### Ethics approval and consent to participate

The study protocol was approved by the Ethics Committee of the University of Electronic Science and Technology of China (approval no. 31682). For participants younger than 18 years, written informed consent was obtained from a parent or legal guardian, together with written informed assent from the adolescent. Participants aged 18 years provided written informed consent independently.

### Consent for publication

Not applicable.

### Availability of data and materials

The de-identified datasets generated and/or analysed during the current study are not publicly available because of ethical restrictions and the sensitive nature of adolescent psychiatric data. They may be made available by the corresponding author upon reasonable request and subject to applicable institutional and ethical approvals.

## Authors’ contributions

P.W.: Conceptualization, Methodology, Formal analysis, Data curation, Visualization, Project administration, Writing–original draft, and Writing–review & editing; P.J.W. and Y.Z.: Investigation, participant recruitment, clinical assessments, data collection, Data curation, Project administration, and Writing–review & editing; X.W.: Validation, and Writing–review & editing; C.L.: Methodology, Formal analysis, Data curation, Validation, and Writing–review & editing; Y.H.: Methodology, Supervision, Resources, Project administration; M.M.: Conceptualization, Methodology, Supervision, Funding acquisition, and Writing–review & editing. All authors contributed to the interpretation of the findings, critically reviewed the manuscript, read and approved the final manuscript, and agreed to be accountable for the work.

## Data Availability

The datasets generated and/or analyzed during the current study are not publicly available because they contain potentially identifiable clinical information from adolescent participants but are available from the corresponding author on reasonable request and subject to approval by the relevant ethics committee.

## 7. Acknowledgements

The authors thank all participants and their families, as well as the clinical and research staff involved in participant recruitment, clinical assessment, and data collection. During the preparation of this manuscript, the authors used ChatGPT and Codex (OpenAI) for English-language refinement, organizational suggestions, and assistance with statistical code development. These tools were not used as autonomous decision-making systems and did not independently determine the study design, analytical strategy, or interpretation of the findings. All AI-assisted text, code, and analytical outputs were critically reviewed and verified by the authors, who take full responsibility for the accuracy and integrity of the final manuscript.

## 8. Abbreviations

BMI: Body mass index
BSSI: Beck Scale for Suicide Ideation
CD-RISC: Connor–Davidson Resilience Scale
CDI: Children’s Depression Inventory
CTQ: Childhood Trauma Questionnaire
DSM-5: Diagnostic and Statistical Manual of Mental Disorders (Fifth Edition)
FAD: Family Assessment Device
HAMD-24: 24-item Hamilton Depression Rating Scale
HAMA-14: 14-item Hamilton Anxiety Rating Scale
HC: Healthy controls
HTMT: Heterotrait–monotrait ratio
MDD: Major depressive disorder
NSSI: Non-suicidal self-injury
OSI: Ottawa Self-Injury Inventory
PCRT: Parent–Child Relationship Diagnostic Test
PCA: Principal Component Analysis
PLS-SEM: Partial least squares structural equation modeling
RRS: Ruminative Responses Scale
SI: Suicidal ideation
SNAP-IV: Swanson, Nolan, and Pelham Rating Scale-IV
SRMR: Standardized root mean square residual

