## Supplementary Material for "Family-related experiences, childhood trauma, and cognitive-emotional correlates across psychopathological domains in adolescent depression"

Table S1. Additional clinical characteristics of the MDD sample

Table S2. Spearman correlations among domain scores

Table S3. Measurement and discriminant validity of latent constructs

Table S4. Structural model quality and predictive performance of the PLS-SEM model for the combined sample

Table S5. Significant standardized specific indirect effects in the PLS-SEM model for the combined sample

Table S6. Sensitivity analysis: comparison between combined-sample and MDD-only models

Table S7. Covariate-adjusted sensitivity analysis

Table S1: Additional clinical characteristics of the MDD sample

| Characteristic | MDD (n=80) |
|---|---|
| HAMD score | 27 [23–33] |
| HAMA score | 22 [17–25] |
| PHQ-15 score | 16 [12–20] |
| **Depressive episodes, n (%)** | |
| First episode | 60 (75.0) |
| Recurrent episode | 20 (25.0) |
| **Depression subtype, n (%)** | |
| Unipolar depression | 70 (87.5) |
| Mixed features | 10 (12.5) |
| **Psychotic features, n (%)** | |
| Non-psychotic | 54 (67.5) |
| Psychotic depression | 26 (32.5) |
| **Drug-naïve status, n (%)** | |
| Yes | 16 (20.0) |
| No | 64 (80.0) |
| **Lifetime suicidal behavior, n (%)** | |
| Absent | 7 (8.8) |
| Suicidal ideation only | 37 (46.3) |
| Suicide plan | 4 (5.0) |
| Suicide attempt | 32 (40.0) |

Table S2: Spearman correlations among domain scores

| Variable | 1 | 2 | 3 | 4 | 5 | 6 | 7 | 8 | 9 |
|---|---|---|---|---|---|---|---|---|---|
| 1. FAD | – | | | | | | | | |
| 2. PCRT | 0.548*** | – | | | | | | | |
| 3. CTQ | 0.743*** | 0.672*** | – | | | | | | |
| 4. RRS | 0.504*** | 0.469*** | 0.634*** | – | | | | | |
| 5. Affective distress | 0.620*** | 0.521*** | 0.705*** | 0.851*** | – | | | | |
| 6. SI | 0.602*** | 0.472*** | 0.664*** | 0.770*** | 0.808*** | – | | | |
| 7. NSSI | 0.451*** | 0.448*** | 0.590*** | 0.710*** | 0.766*** | 0.741*** | – | | |
| 8. SNAP | 0.583*** | 0.587*** | 0.698*** | 0.624*** | 0.716*** | 0.590*** | 0.558*** | – | |
| 9. CD-RISC | -0.621*** | -0.431*** | -0.679*** | -0.647*** | -0.824*** | -0.744*** | -0.721*** | -0.648*** | – |

Values represent Spearman's rank correlation coefficients. Pairwise sample sizes ranged from 130 to 133.

$^{***}p < 0.001$.

FAD = Family Assessment Device composite score; PCRT = Parent–Child Relationship Diagnostic Test composite score; CTQ = Childhood Trauma Questionnaire composite score; RRS = Ruminative Responses Scale composite score; SI = suicidal ideation composite score; NSSI = non-suicidal self-injury spectrum score; SNAP = Self-regulation difficulties composite score; CD-RISC = Connor–Davidson Resilience Scale score.

Table S3: Measurement and discriminant validity of latent constructs

**Panel A. Reliability and convergent validity**

| Construct | Cronbach's $\alpha$ | rho_A | CR ($\rho_c$) | AVE |
|---|---|---|---|---|
| Affective distress | .953 | .954 | .977 | .955 |
| Childhood Trauma | .768 | .817 | .852 | .594 |
| Family Dysfunction | .919 | .930 | .936 | .677 |
| Maladaptive Parenting | .837 | .907 | .887 | .666 |
| NSSI Spectrum | .899 | .899 | .929 | .767 |
| Psychological Resilience | .936 | .940 | .959 | .886 |
| Ruminative Thinking | .938 | .940 | .960 | .889 |
| Self-regulation difficulties | .865 | .884 | .917 | .786 |
| Suicidal Ideation | .912 | .918 | .945 | .851 |

**Panel B. Discriminant validity (HTMT)**

| Construct pair | HTMT | 95% CI (Lower) | 95% CI (Upper) |
|---|---|---|---|
| Family Dysfunction – Maladaptive Parenting | .607 | .511 | .719 |
| Family Dysfunction – Childhood Trauma | .796 | .691 | .890 |
| Family Dysfunction – Psychological Resilience | .688 | .562 | .790 |
| Family Dysfunction – Ruminative Thinking | .561 | .432 | .674 |
| Family Dysfunction – Affective Distress | .664 | .542 | .767 |
| Family Dysfunction – NSSI Spectrum | .456 | .308 | .590 |
| Family Dysfunction – Self-regulation difficulties | .623 | .465 | .753 |
| Family Dysfunction – Suicidal Ideation | .630 | .517 | .731 |
| Maladaptive Parenting – Childhood Trauma | .751 | .636 | .864 |
| Maladaptive Parenting – Psychological Resilience | .447 | .301 | .600 |
| Maladaptive Parenting – Ruminative Thinking | .518 | .376 | .645 |
| Maladaptive Parenting – Affective Distress | .575 | .448 | .692 |
| Maladaptive Parenting – NSSI Spectrum | .538 | .387 | .676 |
| Maladaptive Parenting – Self-regulation difficulties | .697 | .575 | .800 |
| Maladaptive Parenting – Suicidal Ideation | .552 | .390 | .707 |
| Childhood Trauma – Psychological Resilience | .748 | .637 | .846 |
| Childhood Trauma – Ruminative Thinking | .722 | .598 | .831 |
| Childhood Trauma – Affective Distress | .791 | .689 | .888 |
| Childhood Trauma – NSSI Spectrum | .720 | .580 | .842 |
| Childhood Trauma – Self-regulation difficulties | .819 | .651 | .952 |
| Childhood Trauma – Suicidal Ideation | .763 | .648 | .868 |
| Psychological Resilience – Ruminative Thinking | .674 | .566 | .769 |
| Psychological Resilience – Affective Distress | .842 | .781 | .893 |
| Psychological Resilience – NSSI Spectrum | .688 | .597 | .777 |
| Psychological Resilience – Self-regulation difficulties | .697 | .582 | .791 |
| Psychological Resilience – Suicidal Ideation | .792 | .723 | .853 |
| Ruminative Thinking – Affective Distress | .909 | .864 | .948 |
| Ruminative Thinking – NSSI Spectrum | .717 | .621 | .800 |
| Ruminative Thinking – Self-regulation difficulties | .709 | .580 | .817 |
| Ruminative Thinking – Suicidal Ideation | .848 | .765 | .915 |
| NSSI Spectrum – Affective Distress | .776 | .692 | .853 |
| NSSI Spectrum – Self-regulation difficulties | .689 | .554 | .794 |
| NSSI Spectrum – Suicidal Ideation | .777 | .693 | .855 |
| Self-regulation difficulties – Affective Distress | .789 | .692 | .871 |
| Self-regulation difficulties – Suicidal Ideation | .693 | .561 | .812 |
| Suicidal Ideation – Affective Distress | .901 | .851 | .943 |

Note. CR = composite reliability; AVE = average variance extracted; HTMT = heterotrait–monotrait ratio. Values indicate generally acceptable internal consistency, convergent validity, and discriminant validity for all constructs in the final PLS-SEM model.

Table S4: Structural model quality and predictive performance of the PLS-SEM model for the combined sample

**Panel A. Explanatory power**

| Construct | $R^2$ | Adj. $R^2$ | $VIF_{max}$ |
|---|---|---|---|
| Maladaptive Parenting | .360 | .352 | 1.00 |
| Childhood Trauma | .596 | .590 | 1.56 |
| Psychological Resilience | .494 | .486 | 2.01 |
| Ruminative Thinking | .391 | .385 | 1.00 |
| Affective distress | .854 | .851 | 1.85 |
| NSSI Spectrum | .534 | .529 | 2.06 |
| Self-regulation difficulties | .601 | .596 | 1.85 |
| Suicidal Ideation | .709 | .705 | 1.68 |

**Panel B. Predictive performance**

| Construct | $Q^2_{predict}$ | CVPAT loss diff. | $p$ |
|---|---|---|---|
| Maladaptive Parenting | .345 | — | — |
| Childhood Trauma | .490 | — | — |
| Psychological Resilience | .404 | — | — |
| Ruminative Thinking | .265 | — | — |
| Affective distress | .383 | -19.690 | $< .001$ |
| NSSI Spectrum | .160 | — | — |
| Self-regulation difficulties | .316 | — | — |
| Suicidal Ideation | .323 | — | — |

**Panel C. Model quality and collinearity**

| | |
|---|---|
| SRMR (Saturated model) | .082 |
| SRMR (Estimated model) | .099 |
| Maximum VIF | 2.06 |

Note. $R^2$ = explained variance; Adj. $R^2$ = adjusted explained variance; $Q^2_{predict}$ = predictive relevance; CVPAT = cross-validated predictive ability test; VIF = variance inflation factor; SRMR = standardized root mean square residual.

Table S5: Significant standardized specific indirect associations in the PLS-SEM model for the combined sample

| Specific indirect association | $\beta$ | SE | $t$ | $p$ |
|---|---|---|---|---|
| *Indirect associations involving childhood trauma* | | | | |
| Family Dysfunction → Childhood Trauma → NSSI Spectrum | .102 | .047 | 2.169 | .030 |
| Maladaptive Parenting → Childhood Trauma → NSSI Spectrum | .082 | .039 | 2.096 | .036 |
| Family Dysfunction → Maladaptive Parenting → Childhood Trauma → NSSI Spectrum | .049 | .025 | 2.000 | .046 |
| *Indirect associations involving rumination* | | | | |
| Childhood Trauma → Ruminative Thinking → Affective distress | .349 | .042 | 8.413 | < .001 |
| Childhood Trauma → Ruminative Thinking → Suicidal Ideation | .334 | .049 | 6.780 | < .001 |
| Childhood Trauma → Ruminative Thinking → NSSI Spectrum | .223 | .055 | 4.079 | < .001 |
| Childhood Trauma → Ruminative Thinking → Self-regulation difficulties | .175 | .058 | 3.022 | .003 |
| *Indirect associations involving resilience* | | | | |
| Childhood Trauma → Psychological Resilience → Affective distress | .162 | .041 | 3.927 | < .001 |
| Childhood Trauma → Psychological Resilience → Suicidal Ideation | .160 | .045 | 3.570 | < .001 |
| Childhood Trauma → Psychological Resilience → Self-regulation difficulties | .125 | .047 | 2.635 | .008 |
| Childhood Trauma → Psychological Resilience → NSSI Spectrum | .108 | .046 | 2.346 | .019 |
| Family Dysfunction → Psychological Resilience → Affective distress | .142 | .048 | 2.978 | .003 |
| Family Dysfunction → Psychological Resilience → Suicidal Ideation | .140 | .045 | 3.079 | .002 |
| Family Dysfunction → Psychological Resilience → Self-regulation difficulties | .109 | .041 | 2.670 | .008 |
| Family Dysfunction → Psychological Resilience → NSSI Spectrum | .094 | .045 | 2.097 | .036 |
| *Serial indirect associations involving childhood trauma and rumination* | | | | |
| Family Dysfunction → Childhood Trauma → Ruminative Thinking → Affective distress | .167 | .028 | 5.992 | < .001 |
| Family Dysfunction → Childhood Trauma → Ruminative Thinking → Suicidal Ideation | .160 | .030 | 5.323 | < .001 |
| Family Dysfunction → Childhood Trauma → Ruminative Thinking → NSSI Spectrum | .107 | .030 | 3.564 | < .001 |
| Family Dysfunction → Childhood Trauma → Ruminative Thinking → Self-regulation difficulties | .084 | .029 | 2.848 | .004 |
| Maladaptive Parenting → Childhood Trauma → Ruminative Thinking → Affective distress | .134 | .027 | 4.995 | < .001 |
| Maladaptive Parenting → Childhood Trauma → Ruminative Thinking → Suicidal Ideation | .128 | .029 | 4.462 | < .001 |
| Maladaptive Parenting → Childhood Trauma → Ruminative Thinking → NSSI Spectrum | .085 | .025 | 3.485 | < .001 |
| Maladaptive Parenting → Childhood Trauma → Ruminative Thinking → Self-regulation difficulties | .067 | .025 | 2.657 | .008 |
| Family Dysfunction → Maladaptive Parenting → Childhood Trauma → Ruminative Thinking → Affective distress | .080 | .017 | 4.610 | < .001 |
| Family Dysfunction → Maladaptive Parenting → Childhood Trauma → Ruminative Thinking → Suicidal Ideation | .077 | .019 | 4.150 | < .001 |
| Family Dysfunction → Maladaptive Parenting → Childhood Trauma → Ruminative Thinking → NSSI Spectrum | .051 | .015 | 3.385 | .001 |
| Family Dysfunction → Maladaptive Parenting → Childhood Trauma → Ruminative Thinking → Self-regulation difficulties | .040 | .016 | 2.558 | .011 |
| *Serial indirect associations involving childhood trauma and resilience* | | | | |
| Family Dysfunction → Childhood Trauma → Psychological Resilience → Affective distress | .077 | .021 | 3.724 | < .001 |
| Family Dysfunction → Childhood Trauma → Psychological Resilience → Suicidal Ideation | .076 | .022 | 3.412 | .001 |
| Family Dysfunction → Childhood Trauma → Psychological Resilience → Self-regulation difficulties | .060 | .023 | 2.564 | .010 |
| Family Dysfunction → Childhood Trauma → Psychological Resilience → NSSI Spectrum | .051 | .022 | 2.336 | .020 |
| Maladaptive Parenting → Childhood Trauma → Psychological Resilience → Affective distress | .062 | .019 | 3.304 | .001 |
| Maladaptive Parenting → Childhood Trauma → Psychological Resilience → Suicidal Ideation | .061 | .020 | 3.069 | .002 |

| Specific indirect association | $\beta$ | SE | $t$ | $p$ |
|---|---|---|---|---|
| Maladaptive Parenting → Childhood Trauma → Psychological Resilience → Self-regulation difficulties | .048 | .020 | 2.424 | .015 |
| Maladaptive Parenting → Childhood Trauma → Psychological Resilience → NSSI Spectrum | .041 | .019 | 2.145 | .032 |
| Family Dysfunction → Maladaptive Parenting → Childhood Trauma → Psychological Resilience → Affective distress | .037 | .011 | 3.414 | .001 |
| Family Dysfunction → Maladaptive Parenting → Childhood Trauma → Psychological Resilience → Suicidal Ideation | .037 | .012 | 3.143 | .002 |
| Family Dysfunction → Maladaptive Parenting → Childhood Trauma → Psychological Resilience → Self-regulation difficulties | .029 | .011 | 2.497 | .013 |
| Family Dysfunction → Maladaptive Parenting → Childhood Trauma → Psychological Resilience → NSSI Spectrum | .025 | .011 | 2.207 | .027 |
| *Indirect associations involving maladaptive parenting* | | | | |
| Family Dysfunction → Maladaptive Parenting → Self-regulation difficulties | .211 | .046 | 4.631 | $< .001$ |
| Family Dysfunction → Maladaptive Parenting → Affective distress | .059 | .025 | 2.380 | .017 |

Note. Only statistically significant specific indirect association ($p < .05$) are presented. $\beta$ = standardized specific indirect associations; SE = standard error.

Table S6: Direct associations in the combined-sample and MDD-only PLS-SEM models

| Path | Combined sample $\beta$ ($p$) | MDD-only $\beta$ ($p$) | Interpretation |
|---|---|---|---|
| *Family-related structural associations* | | | |
| Family Dysfunction → Maladaptive Parenting | .600 (< .001) | .557 (< .001) | Similar coefficient |
| Family Dysfunction → Childhood Trauma | .478 (< .001) | .444 (< .001) | Similar coefficient |
| Maladaptive Parenting → Childhood Trauma | .383 (< .001) | .359 (< .001) | Similar coefficient |
| *Associations with rumination and resilience* | | | |
| Childhood Trauma → Ruminative Thinking | .625 (< .001) | .373 (< .001) | Smaller coefficient in MDD-only |
| Childhood Trauma → Psychological Resilience | -.405 (< .001) | -.442 (< .001) | Similar coefficient |
| Family Dysfunction → Psychological Resilience | -.355 (.001) | — | Not retained |
| *Associations of rumination and Resilience with Psychopathological domains* | | | |
| Ruminative Thinking → Affective distress | .559 (< .001) | .450 (< .001) | Smaller coefficient in MDD-only |
| Ruminative Thinking → Suicidal Ideation | .535 (< .001) | .345 (< .001) | Smaller coefficient in MDD-only |
| Ruminative Thinking → NSSI Spectrum | .356 (< .001) | .241 (.011) | Smaller coefficient in MDD-only |
| Ruminative Thinking → Self-regulation difficulties | .280 (.001) | — | Not retained |
| Psychological Resilience → Affective distress | -.399 (< .001) | -.383 (< .001) | Similar coefficient |
| Psychological Resilience → Suicidal Ideation | -.394 (< .001) | -.281 (.003) | Smaller coefficient in MDD-only |
| Psychological Resilience → NSSI Spectrum | -.265 (.006) | -.270 (.005) | Similar coefficient |
| Psychological Resilience → Self-regulation difficulties | -.308 (< .001) | -.259 (.007) | Similar coefficient |
| *Additional direct associations with psychopathological domains* | | | |
| Childhood Trauma → NSSI Spectrum | .214 (.023) | — | Not retained |
| Maladaptive Parenting → Affective distress | .098 (.014) | .277 (.001) | Larger coefficient in MDD-only |
| Maladaptive Parenting → Suicidal Ideation | — | .257 (.013) | Additional association in MDD-only |
| Maladaptive Parenting → NSSI Spectrum | — | .309 (.001) | Additional association in MDD-only |
| Maladaptive Parenting → Self-regulation difficulties | .352 (< .001) | .474 (< .001) | Larger coefficient in MDD-only |

Note. Differences in path coefficients between the combined-sample and MDD-only models are descriptive and were not evaluated using formal between-model coefficient tests. “Not retained” indicates an association retained in the combined-sample model but not in the MDD-only model. “Additional association in MDD-only” indicates an association retained only in the MDD-only model.

Table S7: Covariate-adjusted regression sensitivity analyses using PLS-SEM latent variable scores

| Path | Unadjusted $\beta$ | Adjusted $\beta$ | Adjusted 95% CI | $p$ |
|---|---|---|---|---|
| *Family-related structural associations* | | | | |
| Family dysfunction → Maladaptive parenting | .600 | .633 | [.471, .794] | <.001 |
| Family dysfunction → Childhood trauma | .478 | .420 | [.271, .564] | <.001 |
| Maladaptive parenting → Childhood trauma | .383 | .383 | [.252, .513] | <.001 |
| *Associations with rumination and resilience* | | | | |
| Childhood trauma → Ruminative thinking | .625 | .465 | [.306, .615] | <.001 |
| Family dysfunction → Psychological resilience | -.355 | -.226 | [-.422, -.022] | .029 |
| Childhood trauma → Psychological resilience | -.405 | -.313 | [-.530, -.124] | .001 |
| *Associations of rumination and resilience with psychopathological domains* | | | | |
| Ruminative thinking → Affective distress | .559 | .484 | [.390, .582] | <.001 |
| Ruminative thinking → Suicidal ideation | .535 | .458 | [.334, .594] | <.001 |
| Ruminative thinking → NSSI spectrum | .356 | .309 | [.104, .503] | .004 |
| Ruminative thinking → Self-regulation difficulties | .280 | .278 | [.085, .443] | .004 |
| Psychological resilience → Affective distress | -.399 | -.319 | [-.417, -.218] | <.001 |
| Psychological resilience → Suicidal ideation | -.394 | -.278 | [-.393, -.133] | <.001 |
| Psychological resilience → NSSI spectrum | -.265 | -.229 | [-.432, .017] | .067 |
| Psychological resilience → Self-regulation difficulties | -.308 | -.313 | [-.482, -.144] | <.001 |
| *Additional direct associations with psychopathological domains* | | | | |
| Childhood trauma → NSSI spectrum | .214 | .251 | [.025, .470] | .033 |
| Maladaptive parenting → Affective distress | .098 | .129 | [.044, .205] | .004 |
| Maladaptive parenting → Self-regulation difficulties | .352 | .389 | [.244, .519] | <.001 |

Note. Adjusted models included paternal education, maternal education, primary caregiver structure, parental marital status, and academic pressure. Estimates are standardized coefficients from regression-based sensitivity analyses using PLS latent variable scores; 95% confidence intervals were based on 5,000 bootstrap resamples.
